# Early imaging signs associated with subsequent diagnosis of primary sclerosing cholangitis

**DOI:** 10.64898/2026.09.25.26362274

**Authors:** Quentin Vanderbecq, Pauline Iorio, Pierre-Antoine Soret, Sanaa El Mouhadi, Eleonora De Martin, Lionel Arrivé, Sara Lemoinne, Maïté Lewin, Mathilde Wagner

## Abstract

**BACKGROUND:** Primary sclerosing cholangitis (PSC) is characterized by multifocal biliary strictures on MRI/MRCP. However, we have observed that young PSC patients often displayed a marked intrahepatic bile duct visibility, sometimes with gallbladder enlargement, before developing biliary strictures. This study aimed to evaluate whether this imaging pattern was associated with subsequent diagnosis of PSC.

**PATIENTSAND METHODS:** This retrospective two-center study included patients younger than 21 years who had a baseline liver MRI/MRCP, without biliary strictures. The endpoint was a definite diagnosis of large duct PSC during follow-up. The association of baseline imaging findings with the endpoint was evaluated by multivariable logistic regression, Cox models, and Kaplan-Meier analyses.

**RESULTS:** 94 patients were included: 54% women, mean age of 16, 36% with IBD. Over a mean follow-up of 7.75 years, 21 patients were subsequently diagnosed with large duct PSC after a median time of 5.60 years. Marked intrahepatic bile duct visibility and gallbladder enlargement displayed good performance to predict the subsequent diagnosis of PSC (AUC, 0.77 and 0.69, respectively). The presence of both signs displayed a specificity of 1.00. In multivariable analyses, marked intrahepatic bile duct visibility and gallbladder enlargement were independently associated with subsequent PSC diagnosis (OR, 13.35; 95% CI, 3.00– 59.41; p < 0.001 and OR, 12.36; 95% CI, 1.93–79.09; p = 0.008, respectively) and shorter time to diagnosis (HR, 6.42; 95% CI, 1.99–20.75; p = 0.002 and HR, 3.36; 95% CI, 1.22–9.27; p = 0.019, respectively). Survival analyses confirmed the association between these two imaging signs and subsequent PSC diagnosis (HR, 7.78; 95% CI, 2.62–23.16; p < 0.001 and HR, 3.85; 95% CI, 1.64–9.08; p < 0.001, respectively).

**CONCLUSIONS:** In young patients without biliary strictures, marked intrahepatic bile duct visibility and gallbladder enlargement on MRI/MRCP were strongly associated with subsequent diagnosis of large duct PSC.

**IMPACT AND IMPLICATIONS:** Diagnosis of PSC relies on the presence of multifocal biliary strictures. However, early diagnosis of PSC is crucial since it could trigger earlier and more effective therapeutic interventions.

In young patients without biliary strictures, we identified two imaging signs significantly associated with the subsequent diagnosis of large duct PSC: marked intrahepatic bile duct visibility and gallbladder enlargement. These findings should encourage closer monitoring of young patients who display these imaging patterns.

**HIGHLIGHTS:**

- Marked intrahepatic bile duct visibility can predict the subsequent diagnosis of large duct PSC.
- Gallbladder enlargement can predict the subsequent diagnosis of large duct PSC.
- These two signs were significantly associated with subsequent diagnosis of large duct PSC.

## Introduction

Primary sclerosing cholangitis (PSC) is a rare liver disease characterized by multifocal fibro-inflammatory lesions of the intra- and/or extrahepatic bile ducts. These lesions lead to the formation of multiple biliary strictures and eventual progression to biliary cirrhosis and its complications [1]. PSC is strongly associated with inflammatory bowel disease (IBD), particularly ulcerative colitis, and often follows an insidious course with delayed clinical recognition [2,3]. Although PSC is a heterogeneous disease with variable prognosis, PSC patients are at risk for cirrhosis and cholangiocarcinoma and liver transplant-free survival is estimated at 18-21 years in population-based cohorts [4,5] Magnetic resonance cholangiopancreatography (MRCP), typically combined with liver MRI, is now considered as the first-line imaging modality for the diagnosis of PSC. According to recent international guidelines, MRI/MRCP has demonstrated diagnostic performance comparable to endoscopic retrograde cholangiopancreatography (ERCP) in detecting characteristic bile duct changes in established PSC [6–8]. Its ability to demonstrate multifocal strictures alternating with normal or mildly dilated bile duct segments is central to the diagnosis of PSC [6,9]. However, the interpretation of MRI/MRCP remains challenging and is best performed by radiologists with expertise in PSC [6].

During our multidisciplinary discussions including radiologists, hepatologists and interventional endoscopists, we have observed on several occasions that young patients with PSC displayed an early MRI/MRCP pattern characterized by marked intrahepatic bile duct visibility, sometimes associated with gallbladder distension, before the onset of biliary strictures. Because this pattern was repeatedly identified in children, adolescents, and young adults, the current study was deliberately restricted to patients younger than 21 years in order to investigate this pattern in a more homogeneous population.

In this setting, the purpose of the present study was to evaluate whether marked intrahepatic biliary visibility on MRI/MRCP may represent an early imaging sign associated with subsequent diagnosis of large duct PSC in young patients, and to assess the role of associated gallbladder enlargement and quantitative biliary measurements within this imaging pattern.

## Materials and methods

### Study design

This retrospective multicenter study was conducted in two tertiary referral centers specialized in biliary tract diseases. Patients were identified through a systematic review of institutional radiological databases.

### Inclusion criteria

We included patients younger than 21 years who underwent a liver MRI including MRCP (MRI/MRCP) before 2020 in one of the two participating tertiary referral centers. The MRI/MRCP was required to be technically sufficient for assessment of the biliary tree.

### Exclusion criteria

Exclusion criteria included the presence of biliary strictures on baseline MRI/MRCP, prior liver transplantation or hepatic surgery, low phospholipid-associated cholelithiasis (LPAC) syndrome, sickle cell disease, MRI/MRCP examinations deemed non-analyzable for the biliary tract, biliary stones outside the gallbladder, biliary tract malformation, and other conditions considered incompatible with the study objective. For the final analysis, patients from the first analytic cohort were secondarily excluded when follow-up was shorter than 5 years and no alternative diagnosis explaining the MRI/MRCP indication was retained; isolated IBD was not considered sufficient to explain the MRI indication.

### Patient selection

In Center 1, 218 patients were screened. A total of 139 patients were not included because of biliary strictures (n=73), prior liver transplantation/hepatic surgery (n=12), sickle cell disease (n=4), biliary stones visible on MRI/MRCP (n=21), LPAC syndrome (n=7), biliary tract malformation (n=10), or non-analyzable MRI/MRCP (n=12). Thus, 79 patients from Center 1 entered the first analytic cohort.

In Center 2, 158 patients were screened. Exclusions were prior liver transplantation/hepatic surgery (n=69), biliary strictures (n=25), biliary tract malformation (n=21), sickle cell disease (n=6), and LPAC syndrome (n=1), poor MRI/MRCP quality (n=4), common biliary lithiasis without LPAC (n=3), and other causes (n=8). A total of 21 patients from Center 2 were ultimately retained for analysis.

The first analytic cohort comprised 100 patients. Six patients were then excluded from the final analysis because follow-up was shorter than 5 years and no diagnosis explaining the MRI/MRCP indication was retained. The final analytic cohort comprised 94 patients, including 21 with subsequent diagnosis of PSC and 73 without subsequent diagnosis of PSC (Figure 1).

**Figure 1.**
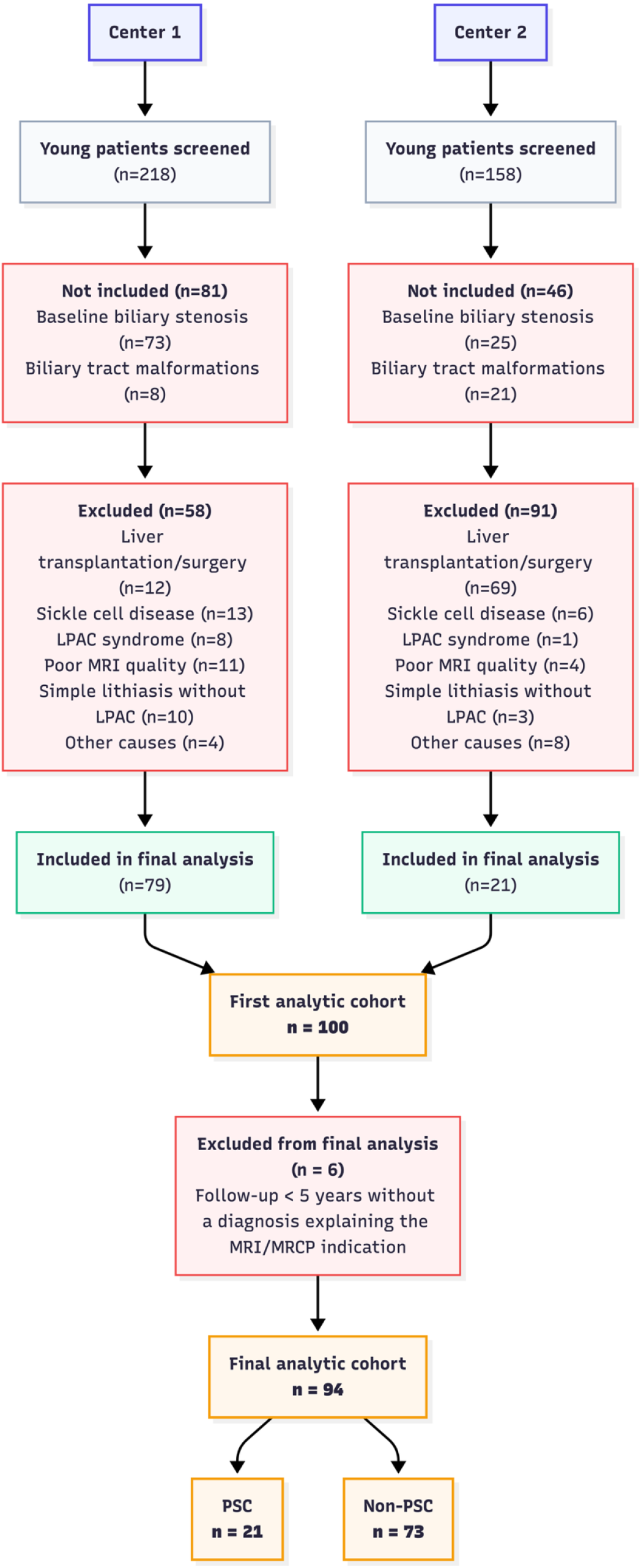
Flowchart of patient selection and construction of the final analytic cohort.

### Baseline assessment and definitions

Baseline was defined by the date of the first MRI/MRCP. At baseline, clinical characteristics (sex, age, presence of IBD) and imaging findings were recorded. The primary diagnostic criterion evaluated in this study was marked intrahepatic bile duct visibility. This was defined as the visualization of the bile ducts beyond the second-order intrahepatic branches on MRCP images, or, alternatively, as the visualization of intrahepatic bile ducts on T2-weighted, or post-contrast T1-weighted images. The secondary imaging criterion was gallbladder distension, which was assessed on MRI and defined as a maximal short-axis gallbladder diameter greater than 30 mm. This threshold was chosen because it corresponded with the qualitative assessment of radiologists for an enlarged gallbladder and was used in previously published works [10]. Additional recorded MRCP/MRI findings included liver dysmorphia (assessed visually by the radiologists and defined as abnormal liver morphology including lobar redistribution, segmental atrophy or hypertrophy, and surface irregularity. In doubtful cases, caudate lobe hypertrophy was assessed using the segment I/right liver lobe ratio, with a ratio >0.65 [11]), splenomegaly (defined as ≥13 cm in the largest axis[12]) portal hypertension (defined by the presence of portosystemic collateral vessels and/or ascites on MRI), bile duct strictures, other biliary tract abnormality. We also reported MRI/MRCP quality (classified as good/excellent, fair or poor [9]), and availability of gadolinium-enhanced or diffusion-weighted sequences.

Quantitative measurements were independently performed by two radiologists and averaged. Biliary measurements included the proximal common bile duct diameter, measured above the cystic duct insertion; the distal common bile duct diameter, measured below the cystic duct insertion; and the right and left hepatic duct diameters. Gallbladder long-axis and short-axis diameters and spleen volume were also recorded.

The endpoint was a definite diagnosis of large duct PSC during follow-up. Definite diagnosis of large duct PSC was made by expert multidisciplinary assessment, in accordance with international guidelines, notably based on the presence of multifocal bile strictures and exclusion of causes of secondary sclerosing cholangitis [8,13]. Patients who were diagnosed for small-duct PSC on liver histology after the baseline MRI/MRCP were not considered to reach the endpoint.

### Imaging analysis

All included MRI/MRCP examinations were independently reviewed by two senior radiologists with 4 and 5 years of experience in hepatobiliary imaging, respectively, both practicing in specialized centers for biliary tract pathologies. In case of disagreement between the two readers, a consensus review was conducted with the participation of a third senior radiologist with more than 10 years of experience in biliary tract imaging, and final decisions were reached by consensus. The first radiologist reviewed all initially selected MRI/MRCP examinations to confirm eligibility according to the predefined criteria. In cases where exclusion was not unequivocal, the same review process was applied, involving an independent assessment by the second annotator and, if necessary, adjudication was made by the third radiologist.

### Ethical approval

Ethical approval for this retrospective observational study was granted by the CERIM (reference CRM-2512-522). The study was conducted in accordance with the Declaration of Helsinki and French MR-004 regulations. As the study used anonymized data collected during routine care, the requirement for individual informed consent was waived in accordance with French law; patients were informed that their data could be used for research purposes.

### Statistical analysis

Statistical analyses were performed using Python, including the scikit-learn library [14]. Continuous variables were expressed as mean (standard deviation) and compared using Mann–Whitney U tests. Categorical variables were expressed as number and percentage and compared using Fisher’s exact tests. Diagnostic performance was summarized using area under the receiver operating characteristic curve (AUC), sensitivity, specificity, positive predictive value (PPV), and negative predictive value (NPV). Multivariable logistic regression assessed associations with final PSC status. Time-to-event analyses used Kaplan-Meier curves, log-rank tests, and Cox proportional hazards models.

Multivariable models included marked bile duct visibility, gallbladder enlargement, age at first MRI/MRCP, IBD, and MRI/MRCP quality. Inter-reader reproducibility was assessed using observed agreement and Cohen’s kappa.

## Results

### Study population

The study cohort included 94 young patients with a baseline MRI/MRCP without any biliary stricture. Among the 94 patients, during the follow-up, 21 patients were subsequently diagnosed with large duct PSC (referred to as the PSC group) (Figure 2) and 73 were not (referred to as the non-PSC group). In patients who developed large duct PSC, the median time from baseline to PSC diagnosis was 5.60 years [IQR, 4.41-6.76]. At baseline, mean age was 16.70 years (SD, 3.49), and 51/94 patients (54.3%) were female. IBD was present in 34/94 patients (36.2%): 12/21 (57.1%) in the PSC group and 22/73 (30.1%) in the non-PSC group (p = 0.038). Baseline MRI/MRCP examinations were performed between October 2006 and October 2020. Mean follow-up duration was 7.75 years (SD, 3.80) overall, 9.11 years (SD, 3.05) in the PSC group, and 7.35 years (SD, 3.92) in the non-PSC group (p = 0.06). (Table 1) All final etiologies are presented in Supplementary Table 1. Etiologies classified as other included specific conditions that could not be consistently reassigned to the predefined categories.

**Table 1.**
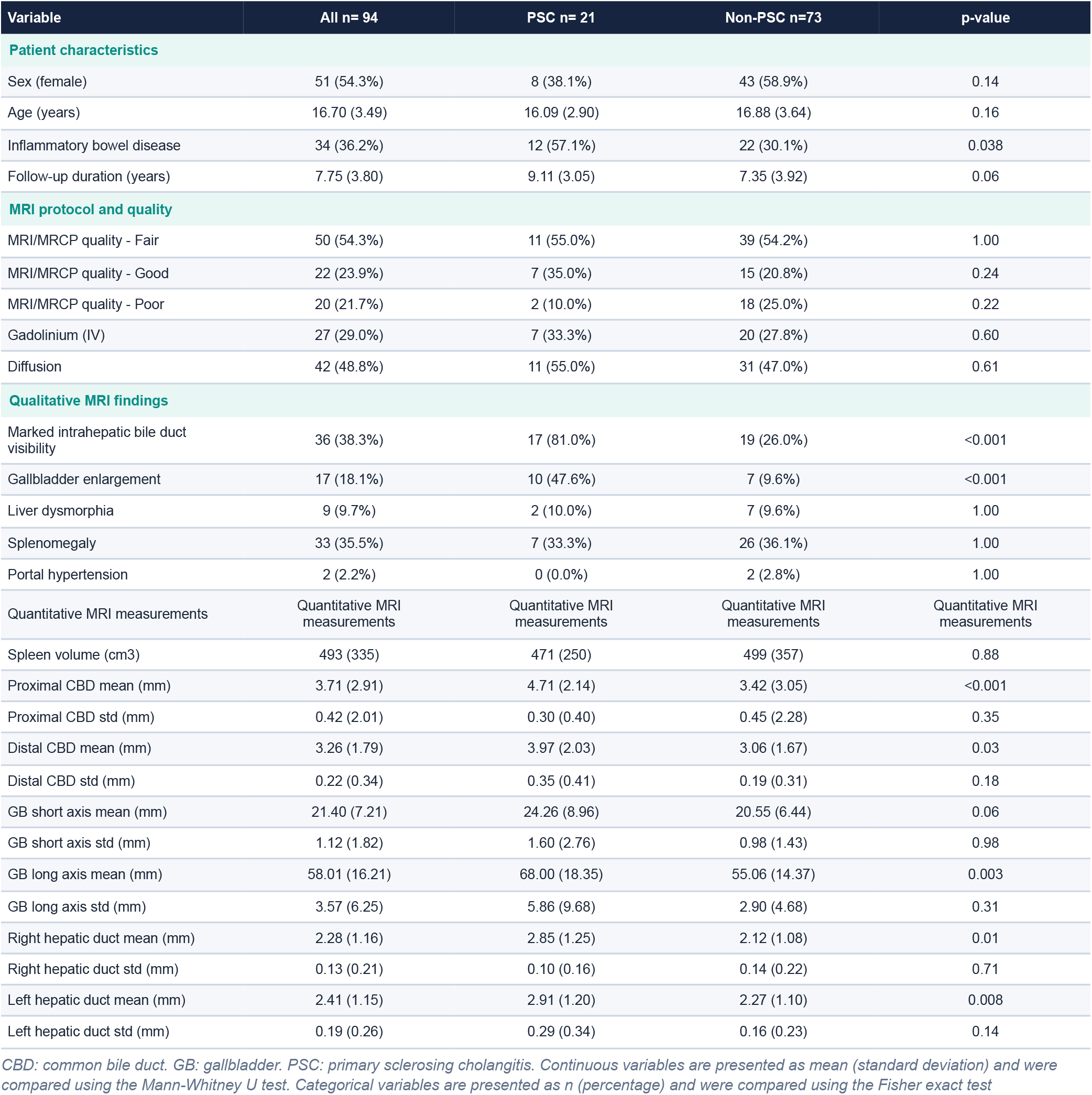
Baseline characteristics and imaging findings.

| Variable | All n= 94 | PSC n= 21 | Non-PSC n=73 | p-value |
| --- | --- | --- | --- | --- |
| **Patient characteristics** |  |  |  |  |
| Sex (female) | 51 (54.3%) | 8 (38.1%) | 43 (58.9%) | 0.14 |
| Age (years) | 16.70 (3.49) | 16.09 (2.90) | 16.88 (3.64) | 0.16 |
| Inflammatory bowel disease | 34 (36.2%) | 12 (57.1%) | 22 (30.1%) | 0.038 |
| Follow-up duration (years) | 7.75 (3.80) | 9.11 (3.05) | 7.35 (3.92) | 0.06 |
| **MRI protocol and quality** |  |  |  |  |
| MRI/MRCP quality - Fair | 50 (54.3%) | 11 (55.0%) | 39 (54.2%) | 1.00 |
| MRI/MRCP quality - Good | 22 (23.9%) | 7 (35.0%) | 15 (20.8%) | 0.24 |
| MRI/MRCP quality - Poor | 20 (21.7%) | 2 (10.0%) | 18 (25.0%) | 0.22 |
| Gadolinium (IV) | 27 (29.0%) | 7 (33.3%) | 20 (27.8%) | 0.60 |
| Diffusion | 42 (48.8%) | 11 (55.0%) | 31 (47.0%) | 0.61 |
| **Qualitative MRI findings** |  |  |  |  |
| Marked intrahepatic bile duct visibility | 36 (38.3%) | 17 (81.0%) | 19 (26.0%) | <0.001 |
| Gallbladder enlargement | 17 (18.1%) | 10 (47.6%) | 7 (9.6%) | <0.001 |
| Liver dysmorphia | 9 (9.7%) | 2 (10.0%) | 7 (9.6%) | 1.00 |
| Splenomegaly | 33 (35.5%) | 7 (33.3%) | 26 (36.1%) | 1.00 |
| Portal hypertension | 2 (2.2%) | 0 (0.0%) | 2 (2.8%) | 1.00 |
| Quantitative MRI measurements | Quantitative MRI measurements | Quantitative MRI measurements | Quantitative MRI measurements | Quantitative MRI measurements |
| Spleen volume (cm3) | 493 (335) | 471 (250) | 499 (357) | 0.88 |
| Proximal CBD mean (mm) | 3.71 (2.91) | 4.71 (2.14) | 3.42 (3.05) | <0.001 |
| Proximal CBD std (mm) | 0.42 (2.01) | 0.30 (0.40) | 0.45 (2.28) | 0.35 |
| Distal CBD mean (mm) | 3.26 (1.79) | 3.97 (2.03) | 3.06 (1.67) | 0.03 |
| Distal CBD std (mm) | 0.22 (0.34) | 0.35 (0.41) | 0.19 (0.31) | 0.18 |
| GB short axis mean (mm) | 21.40 (7.21) | 24.26 (8.96) | 20.55 (6.44) | 0.06 |
| GB short axis std (mm) | 1.12 (1.82) | 1.60 (2.76) | 0.98 (1.43) | 0.98 |
| GB long axis mean (mm) | 58.01 (16.21) | 68.00 (18.35) | 55.06 (14.37) | 0.003 |
| GB long axis std (mm) | 3.57 (6.25) | 5.86 (9.68) | 2.90 (4.68) | 0.31 |
| Right hepatic duct mean (mm) | 2.28 (1.16) | 2.85 (1.25) | 2.12 (1.08) | 0.01 |
| Right hepatic duct std (mm) | 0.13 (0.21) | 0.10 (0.16) | 0.14 (0.22) | 0.71 |
| Left hepatic duct mean (mm) | 2.41 (1.15) | 2.91 (1.20) | 2.27 (1.10) | 0.008 |
| Left hepatic duct std (mm) | 0.19 (0.26) | 0.29 (0.34) | 0.16 (0.23) | 0.14 |

CBD: common bile duct. GB: gallbladder. PSC: primary sclerosing cholangitis. Continuous variables are presented as mean (standard deviation) and were compared using the Mann-Whitney U test. Categorical variables are presented as n (percentage) and were compared using the Fisher exact test

**Figure 2.**
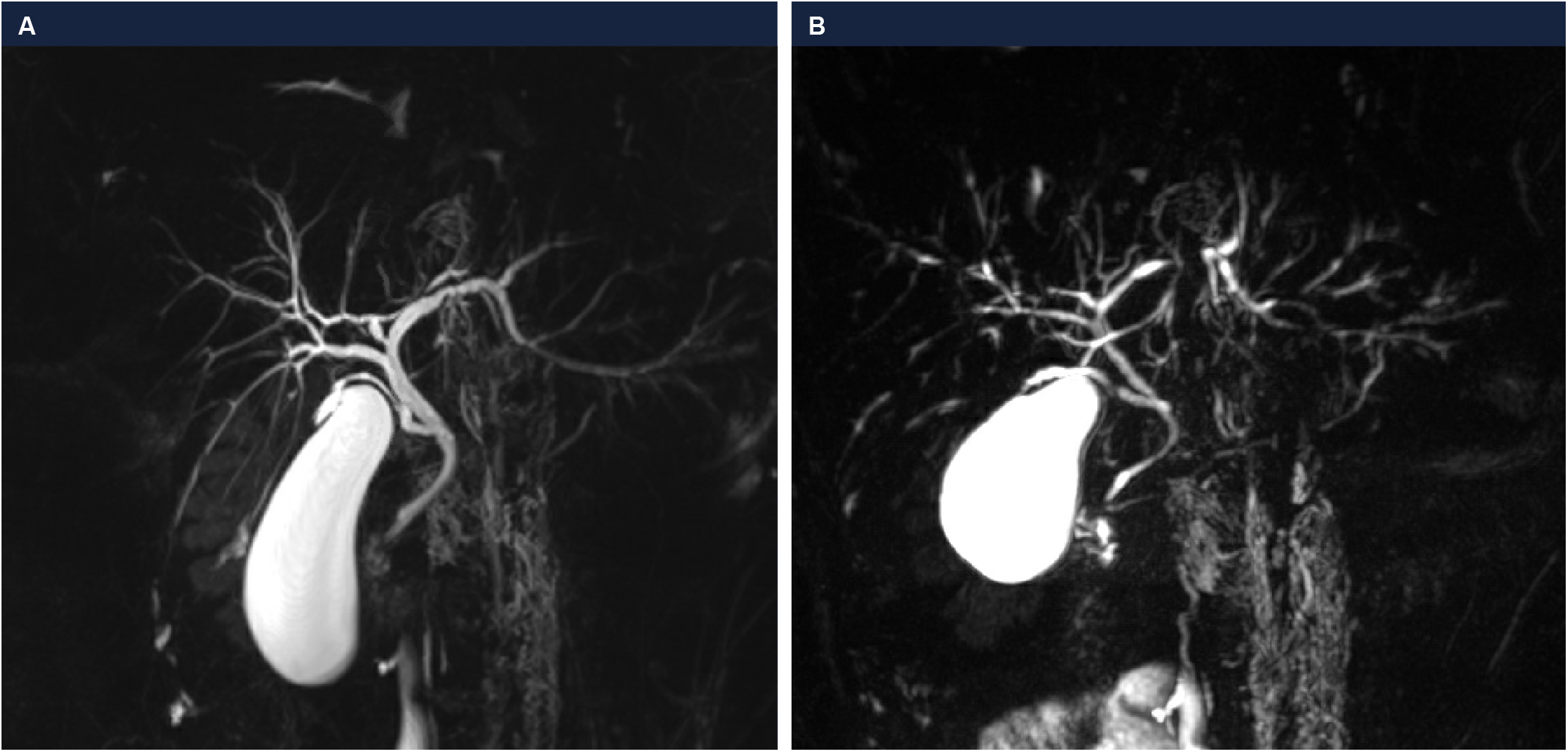
Representative MRI/MRCP findings in a young patient with marked intrahepatic bile duct visibility and gallbladder enlargement who subsequently developed PSC. (a) A male patient aged 15–19 years with ulcerative colitis and isolated elevated alanine aminotransferase levels underwent baseline MRI/MRCP, which demonstrated marked visibility of the intrahepatic bile ducts beyond the second-order branches, associated with gallbladder enlargement but no definite biliary stricture. (b) Follow-up MRCP performed several years later showing typical features of large duct primary sclerosing cholangitis, with multiple intrahepatic biliary strictures

### Performance of qualitative imaging findings

Marked intrahepatic bile duct visibility was present in 36/94 patients (38.3%) overall. It was significantly more frequent in the PSC group (17/21 patients, 81.0%) than in the non-PSC group (19/73 patients, 26.0%) (p < 0.001). Gallbladder enlargement was present in 17/94 patients (18.1%). It was significantly more frequent in the PSC group (10/21 patients, 47.6%) than in the non-PSC group (7/73 patients, 9.6%) (p < 0.001). The performance of these two signs is presented in Table 2. The combination of both signs was present in 9/94 patients (9.6%), all of whom developed subsequent PSC, yielding specificity and PPV of 1.00, with sensitivity of 0.43 (Table 2). Representative examples of the two imaging signs are shown in Figure 2.

**Table 2.** Performance of qualitative imaging findings to predict subsequent diagnosis of PSC.

| Predictor | AUC | Sensitivity | Specificity | PPV | NPV |
| --- | --- | --- | --- | --- | --- |
| Marked intrahepatic bile duct visibility | 0.77 | 0.81 | 0.74 | 0.47 | 0.93 |
| Gallbladder enlargement | 0.69 | 0.48 | 0.90 | 0.59 | 0.86 |
| Marked intrahepatic bile duct visibility + gallbladder enlargement | 0.71 | 0.43 | 1.00 | 1.00 | 0.86 |

AUC: area under the receiver operating characteristic curve. NPV: negative predictive value. PPV: positive predictive value.

### Performance of quantitative imaging findings

Mean proximal common bile duct diameter was significantly larger in the PSC group than in the non-PSC group (4.71 vs 3.42 mm; p < 0.001), as were distal common bile duct diameter (3.97 vs 3.06 mm; p = 0.03), right hepatic duct diameter (2.85 vs 2.12 mm; p = 0.01), left hepatic duct diameter (2.91 vs 2.27 mm; p = 0.008), and gallbladder long-axis diameter (68.00 vs 55.06 mm; p = 0.003).

Exploratory ROC analysis identified Youden-optimized thresholds of 2.5 mm for proximal common bile duct diameter, 3.5 mm for distal common bile duct diameter, 3.0 mm for right hepatic duct diameter, and 3.0 mm for left hepatic duct diameter.

### Findings associated with subsequent PSC diagnosis

In multivariable logistic regression, marked intrahepatic bile duct visibility and gallbladder enlargement were independently associated with subsequent diagnosis of PSC. In the model including both imaging signs, age at first MRI/MRCP, IBD, and MRI/MRCP quality, marked intrahepatic bile duct visibility had an adjusted odds ratio of 13.35 (95% CI, 3.00–59.41; p < 0.001), and gallbladder enlargement had an adjusted odds ratio of 12.36 (95% CI, 1.93–79.09; p = 0.008) (Table 3). IBD was also associated with subsequent diagnosis of PSC (adjusted OR, 4.34; 95% CI, 1.03–18.29; p = 0.046); age and MRI/MRCP quality were not significantly associated.

**Table 3.** Adjusted associations with subsequent diagnosis of PSC and time to PSC diagnosis.

| Variable | Logistic model OR (95% CI) | Logistic p-value | Cox model HR (95% CI) | Cox p-value |
| --- | --- | --- | --- | --- |
| Marked intrahepatic bile duct visibility | 13.35 (3.00–59.41) | <0.001 | 6.42 (1.99–20.75) | 0.002 |
| Gallbladder enlargement | 12.36 (1.93–79.09) | 0.008 | 3.36 (1.22–9.27) | 0.019 |
| Age at first MRI/MRCP | 0.93 (0.76–1.14) | 0.481 | 0.95 (0.81–1.12) | 0.528 |
| Inflammatory bowel disease | 4.34 (1.03–18.29) | 0.046 | 1.80 (0.67–4.82) | 0.245 |
| MRI/MRCP quality, Fair vs Good | 0.27 (0.05–1.37) | 0.115 | 0.56 (0.21–1.46) | 0.232 |
| MRI/MRCP quality, Poor vs Good | 0.13 (0.01–1.39) | 0.091 | 0.41 (0.08–2.07) | 0.283 |

Multivariable logistic and Cox models included marked intrahepatic bile duct visibility, gallbladder enlargement, age at first MRI/MRCP, inflammatory bowel disease, and MRI/MRCP quality. The Cox model used time from baseline MRI/MRCP to PSC diagnosis or last follow-up ( $n = 94$ ; 21 PSC events).

In exploratory multivariable Cox analysis, marked intrahepatic bile duct visibility was associated with shorter time to PSC diagnosis (HR, 6.42; 95% CI, 1.99–20.75; p = 0.002), as was gallbladder enlargement (HR, 3.36; 95% CI, 1.22–9.27; p = 0.019) (Figure 3; Table 3). Univariable time-to-event analyses showed similar associations for marked intrahepatic bile duct visibility (HR, 7.78; 95% CI, 2.62–23.16; log-rank p < 0.001) and gallbladder enlargement (HR, 3.85; 95% CI, 1.64– 9.08; log-rank p < 0.001) (Figure 4).

**Figure 3.**
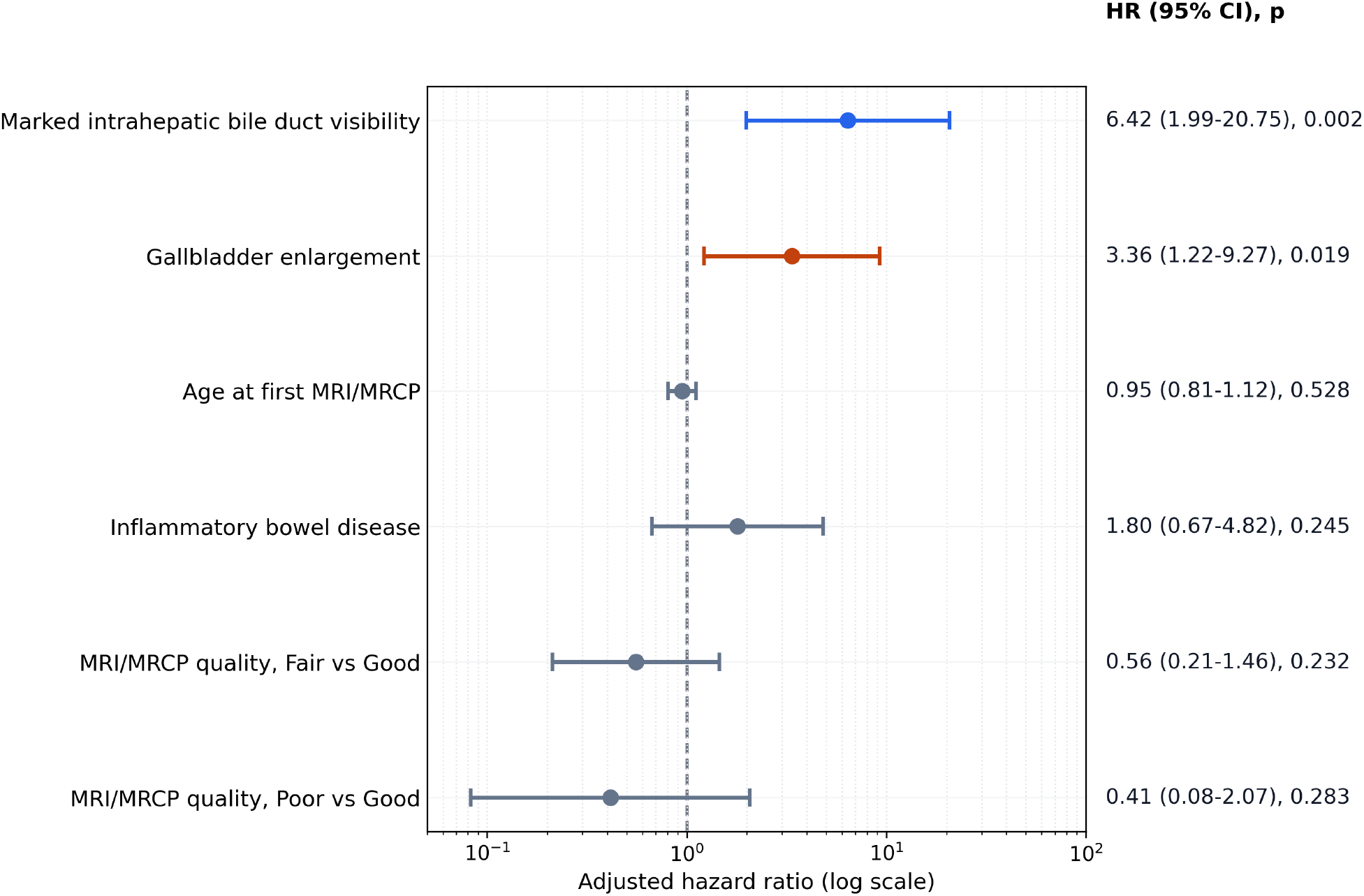
Adjusted Cox proportional hazards model for characteristics associated with subsequent diagnosis of PSC.

**Figure 4.**
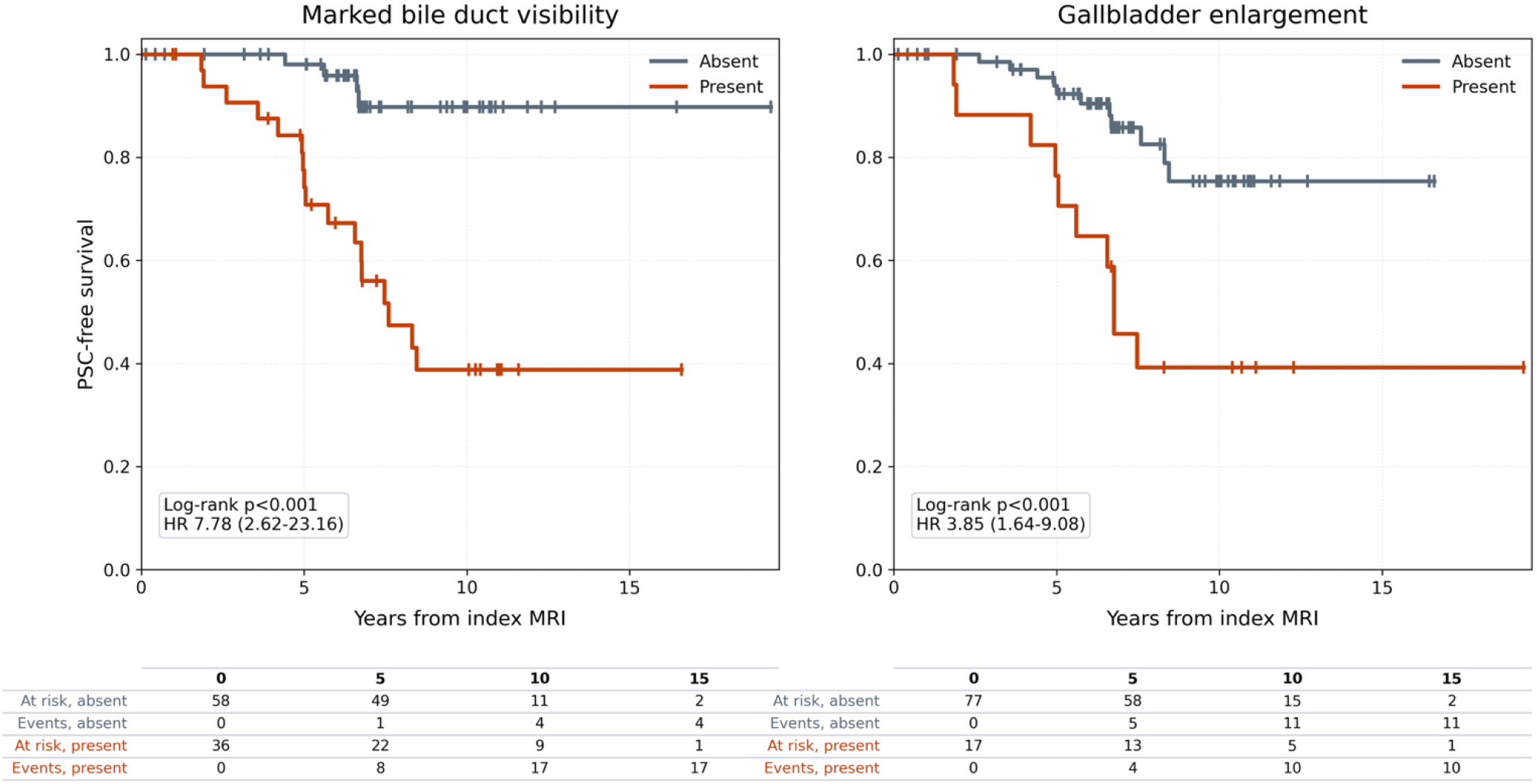
Kaplan-Meier curves showing PSC-free survival according to baseline MRI/MRCP findings. Left: comparison between patients with and without marked visibility of the intrahepatic bile ducts at baseline. Right: comparison between patients with and without gallbladder enlargement at baseline. Censoring ticks indicate patients without PSC diagnosis at last follow-up.

### Inter-reader reproducibility

Inter-reader reproducibility was moderate for marked intrahepatic bile duct visibility and substantial for gallbladder enlargement. (Table 4).

**Table 4.** Inter-reader reproducibility of the qualitative imaging signs.

| Imaging feature | Observed agreement | Cohen's kappa | Interpretation |
| --- | --- | --- | --- |
| Marked intrahepatic bile duct visibility | 74.7% | 0.499 | Moderate |
| Gallbladder enlargement | 90.7% | 0.758 | Substantial |

## Discussion

In this cohort of young patients with normal baseline MRI/MRCP without any biliary strictures, marked visualization of the intrahepatic bile ducts and gallbladder enlargement were independently and strongly associated with subsequent diagnosis of large duct PSC in both logistic and time-to-event analyses. Compared to the marked intrahepatic bile duct visibility, gallbladder enlargement showed a different predictive performance, with lower sensitivity but higher specificity. The association of these two signs showed a good performance to predict subsequent diagnosis of PSC with a specificity of 100% and a PPV of 100%. In addition, quantitative measurements of the common bile duct, right and left hepatic ducts, and gallbladder long axis were significantly higher in patients who were subsequently diagnosed with PSC, supporting the results obtained for the qualitative imaging findings.

Marked intrahepatic bile duct visibility can be considered as a subtle dilatation of the intrahepatic biliary tree. In PSC, dilatations of the bile ducts are usually associated with bile duct strictures and are thought to be the mechanical consequences of these strictures. But in this cohort of young patients with baseline MRI/MRCP without any strictures, the mechanism of the dilatation of the biliary tree is unknown. We hypothesize the implication of early remodeling of the cells of the bile duct wall. Both cholangiocytes and portal fibroblasts could undergo pro-inflammatory changes that impair their functions to maintain homeostasis conditions such as tone of bile ducts. Previously, we reported a subpopulation of PSC patients with a radiological phenotype of intrahepatic cystic biliary dilatation[15]. These patients displayed important dilatation of intrahepatic biliary ducts without necessarily having biliary strictures, suggesting a mechanism of destruction of the bile duct wall. Interestingly, patients with this specific imaging pattern were characterized by a younger age [15].

The association between gallbladder enlargement and PSC is also biologically and radiologically plausible. Increased fasting gallbladder volume has previously been reported in PSC compared with healthy controls and patients with primary biliary cholangitis, with the authors proposing gallbladder volume as a potential non-invasive clue in patients suspected of PSC [16]. Subsequent work found increased fasting and postprandial gallbladder volumes in PSC, without clear evidence of impaired ejection fraction, and suggested that gallbladder mucosal dysfunction or chronic cholecystitis could contribute to the enlargement [17]. Gallbladder abnormalities are common in PSC more broadly [18], and recent work has also raised the possibility that gallbladder enlargement could impact bile acid metabolism and have a protective role in PSC patients [19]. In our cohort, gallbladder enlargement was not sufficiently sensitive to serve as a screening sign, but its high specificity and independent association with PSC suggest that it may be a useful complementary marker when observed with marked bile duct visibility.

Because MRI/MRCP is already the reference non-invasive imaging modality for PSC diagnosis, our quantitative findings should mainly be interpreted as exploratory markers of earlier disease. Existing MRI/MRCP scores, such as ANALI and DiStrict, have been reported to have a prognostic value in patients with established PSC [20–22]. Quantitative MRCP tools, which have also been reported to have a prognostic value in PSC patients, may help define more reproducible thresholds for subtle ductal enlargement before the development of biliary strictures [23,24]. In our cohort, manually performed measurements showed statistically significant differences in bile duct and gallbladder dimensions between groups. However, these differences were submillimeter and are therefore likely to be sensitive to inter-operator variability. Thus, these results should be used as a rationale for future studies using reproducible quantitative MRCP. Early detection of such abnormalities could ultimately help to select patients for closer monitoring and, potentially, for trials of early therapeutic interventions.

This study has several limitations. First, its retrospective design and the inclusion of patients from two tertiary referral centers introduce an inherent selection bias. Therefore, the diagnostic performances reported here should not be directly extrapolated to a screening population. These results are best interpreted as applying to young patients already referred to dedicated hepatobiliary MRI/MRCP in expert settings, rather than to an unselected pediatric or general radiology population. Second, MRI/MRCP examinations were acquired over a long period with heterogeneous protocols across centers. Although all examinations included biliary MR imaging and image quality was assessed, differences in sequence type, spatial resolution, respiratory motion, and biliary tree distension may have influenced duct visibility.

Finally, marked intrahepatic bile duct visibility and gallbladder enlargement remain partly subjective imaging signs with inter-reader variability. However, this inter-reader variability has been reported for all other imaging signs of PSC, even for the presence of high-grade strictures which was associated with a kappa at 0.31 only [25].

Our study also has several strengths. Although its size is limited, our study population is very homogeneous with a significant duration of follow-up. MRI/MRCP assessment was thorough and was performed by two radiologists with expertise in PSC. To our knowledge, this study is the first to identify early imaging signs associated with the subsequent diagnosis of large duct PSC and including patients who did not develop PSC during follow-up. The concept of early or subclinical PSC is supported by several lines of evidence. MR cholangiography screening studies in patients with IBD have shown that PSC-like lesions may be more frequent than clinically recognized PSC, suggesting that biliary disease can remain unrecognized for prolonged periods [26]. Recently, the importance of improving the early imaging diagnosis before the development of biliary strictures has been reported [27]. Indeed, we can assume that treatment may be more effective when initiated earlier, particularly before fibrotic biliary strictures develop. This is particularly relevant given the emerging therapeutic options being assessed in PSC patients [28–30].

## Conclusion

In young patients without biliary strictures, marked intrahepatic bile duct visibility and gallbladder enlargement on MRI/MRCP were strongly associated with subsequent diagnosis of large duct PSC. These findings should encourage closer monitoring of patients who display these imaging patterns.

## FUNDING

The authors received no specific funding for this work.

### AUTHOR CONTRIBUTIONS

Quentin Vanderbecq, Mathilde Wagner, Maïté Lewin, and Lionel Arrivé contributed to the conception and design of the study. Quentin Vanderbecq acquired and curated the data, performed the imaging review and statistical analyses, and drafted the manuscript. Quentin Vanderbecq, Mathilde Wagner, Maïté Lewin, Sara Lemoinne, and Lionel Arrivé contributed to the analysis and interpretation of the data. Pauline Iorio and Sanaa El Mouhadi contributed to data acquisition through imaging review and critically revised the manuscript for important intellectual content. Pierre-Antoine Soret, Eleonora De Martin, and Sara Lemoinne contributed to data interpretation, including clinical interpretation and patient classification, and critically revised the manuscript for important intellectual content. All authors contributed to revising the manuscript critically for important intellectual content and approved the final version of the manuscript for submission.

### DATA AVAILABILITY

The data are available from the corresponding author upon reasonable request.

### ARTIFICIAL INTELLIGENCE DISCLOSURE

Artificial intelligence–assisted tools (ChatGPT, OpenAI and Claude, Anthropic) were used to support language editing and clarity of the manuscript. The authors reviewed, edited, and approved all content and take full responsibility for the accuracy and integrity of the work.

#### ACKNOWLEDGMENTS

Q.V. acknowledges support from the Inria–AP-HP Hosting Position Program (“Poste d’Accueil Inria–AP-HP”).

#### Abbreviations

AUC: area under the receiver operating characteristic curve
CI: confidence interval
ERCP: endoscopic retrograde cholangiopancreatography
IBD: inflammatory bowel disease
LPAC: low phospholipid-associated cholelithiasis
MRI: magnetic resonance imaging
OR: odds ratio
PSC: primary sclerosing cholangitis
SD: standard deviation
CBD: common bile duct
GB: gallbladder
HR: hazard ratio
IQR: interquartile range
MRCP: magnetic resonance cholangiopancreatography
NPV: negative predictive value
PPV: positive predictive value
ROC: receiver operating characteristic curve

