## Supplemental Table 1 for "Early imaging signs associated with subsequent diagnosis of primary sclerosing cholangitis"

MEDRXIV PREPRINT · SUPPLEMENTARY MATERIAL

**Supplementary Table S1.** Definite etiologies diagnosed during follow-up.

| Etiologies | All (n=94) | PSC (n=21) | Non-PSC (n=73) | Bile duct visibility + (n=36) | Bile duct visibility - (n=58) | Gallbladder enlargement + (n=17) | Gallbladder enlargement - (n=77) |
|---|---|---|---|---|---|---|---|
| PSC | 21 (22.3%) | 21 (100%) | 0 (0.0%) | 17 (47.2%) | 4 (6.9%) | 10 (58.8%) | 11 (14.3%) |
| Small-duct PSC | 7 (7.4%) | 0 (0.0%) | 7 (9.6%) | 2 (5.6%) | 5 (8.6%) | 2 (11.8%) | 5 (6.5%) |
| Autoimmune hepatitis | 25 (26.6%) | 6 (28.6%) | 19 (26.0%) | 11 (30.6%) | 14 (24.1%) | 6 (35.3%) | 19 (24.7%) |
| Acute pancreatitis | 15 (16.0%) | 0 (0.0%) | 15 (20.5%) | 7 (19.4%) | 8 (13.8%) | 1 (5.9%) | 14 (18.2%) |
| IBD-associated unexplained liver test abnormalities | 7 (7.4%) | 0 (0.0%) | 7 (7.4%) | 1 (2.8%) | 6 (10.3%) | 1 (5.9%) | 6 (7.8%) |
| Metabolic dysfunction-associated steatotic liver disease (MASLD) | 6 (6.4%) | 0 (0.0%) | 6 (8.2%) | 0 (0.0%) | 6 (10.3%) | 0 (0.0%) | 6 (7.8%) |
| Viral hepatitis | 4 (4.3%) | 0 (0.0%) | 4 (5.5%) | 1 (2.8%) | 3 (5.2%) | 0 (0.0%) | 4 (5.2%) |
| Abdominal trauma | 2 (2.1%) | 0 (0.0%) | 2 (2.7%) | 1 (2.8%) | 1 (1.7%) | 0 (0.0%) | 2 (2.6%) |
| Ischemic cholangitis | 2 (2.1%) | 0 (0.0%) | 2 (2.7%) | 0 (0.0%) | 2 (3.4%) | 0 (0.0%) | 2 (2.6%) |
| Drug-induced liver injury | 1 (1.1%) | 0 (0.0%) | 1 (1.4%) | 1 (2.8%) | 0 (0.0%) | 0 (0.0%) | 1 (1.3%) |
| Other systemic condition (e.g., sarcoidosis) | 6 (6.4%) | 0 (0.0%) | 6 (8.2%) | 2 (5.6%) | 4 (6.9%) | 0 (0.0%) | 6 (7.8%) |
| Indeterminate | 8 (8.5%) | 0 (0.0%) | 8 (11.0%) | 0 (0.0%) | 8 (13.8%) | 2 (11.8%) | 6 (7.8%) |
